# COL6A3-derived endotrophin mediates the effect of obesity on coronary artery disease: an integrative proteogenomics analysis

**DOI:** 10.1101/2023.04.19.23288706

**Authors:** Satoshi Yoshiji, Tianyuan Lu, Guillaume Butler-Laporte, Julia Carrasco-Zanini-Sanchez, Yiheng Chen, Kevin Liang, Julian Daniel Sunday Willett, Chen-Yang Su, Shidong Wang, Darin Adra, Yann Ilboudo, Takayoshi Sasako, Vincenzo Forgetta, Yossi Farjoun, Hugo Zeberg, Sirui Zhou, Michael Hultström, Mitchell Machiela, Nicholas J. Wareham, Vincent Mooser, Nicholas J. Timpson, Claudia Langenberg, J. Brent Richards

**Affiliations:** Lady Davis Institute, Jewish General Hospital, McGill University, Montréal, Québec, Canada; Department of Human Genetics, McGill University, Montréal, Québec, Canada; Kyoto-McGill International Collaborative Program in Genomic Medicine, Graduate School of Medicine, Kyoto University, Kyoto, Japan; Japan Society for the Promotion of Science, Japan; Department of Statistical Sciences, University of Toronto, Canada; Prime Sciences, Montréal, Québec, Canada; Department of Epidemiology, Biostatistics and Occupational Health, McGill University, Montréal, Québec, Canada; Wellcome Trust Centre for Human Genetics, University of Oxford, Oxford, UK; MRC Epidemiology Unit, Institute of Metabolic Science, University of Cambridge, Cambridge, UK; Quantitative Life Sciences Program, McGill University, Montréal, Québec, Canada; SomaLogic, Boulder, Colorado, USA; Fulcrum Genomics, Colorado, USA; Department of Physiology and Pharmacology, Karolinska Institutet, Stockholm, Sweden; Max Planck Institute for Evolutionary Anthropology, Leipzig, Germany; McGill Genome Centre, McGill University, Montréal, Québec, Canada; Anaesthesiology and Intensive Care Medicine, Department of Surgical Sciences, Uppsala University, Uppsala, Sweden; Integrative Physiology, Department of Medical Cell Biology, Uppsala University, Uppsala, Sweden; Division of Cancer Epidemiology and Genetics, National Cancer Institute, Rockville, MD, USA; Integrative Epidemiology Unit, University of Bristol, Bristol, UK; Population Health Sciences, Bristol Medical School, University of Bristol, Bristol, UK; Computational Medicine, Berlin Institute of Health (BIH) at Charité – Universitätsmedizin Berlin, Germany; Precision Healthcare University Research Institute, Queen Mary University of London, London, UK; Department of Twin Research, King’s College London, London, UK

## Abstract

Obesity strongly increases the risk of cardiometabolic diseases, yet the underlying mediators of this relationship are not fully understood. Given that obesity has broad effects on circulating protein levels, we investigated circulating proteins that mediate the effects of obesity on coronary artery disease (CAD), stroke, and type 2 diabetes—since doing so may prioritize targets for therapeutic intervention. By integrating proteome-wide Mendelian randomization (MR) screening 4,907 plasma proteins, colocalization, and mediation analyses, we identified seven plasma proteins, including collagen type VI α3 (COL6A3). COL6A3 was strongly increased by body mass index (BMI) (*β* = 0.32, 95% CI: 0.26–0.38, *P* = 3.7 × 10^-8^ per s.d. increase in BMI) and increased the risk of CAD (OR = 1.47, 95% CI:1.26–1.70, *P* = 4.5 × 10^-7^ per s.d. increase in COL6A3). Notably, COL6A3 is cleaved at its C-terminus to produce endotrophin, which was found to mediate this effect on CAD. In single-cell RNA sequencing of adipose tissues and coronary arteries, *COL6A3* was highly expressed in cell types involved in metabolic dysfunction and fibrosis. Finally, we found that body fat reduction can reduce plasma levels of COL6A3-derived endotrophin, thereby highlighting a tractable way to modify endotrophin levels. In summary, we provide actionable insights into how circulating proteins mediate the effect of obesity on cardiometabolic diseases and prioritize endotrophin as a potential therapeutic target.

## Background

Over 1.9 billion people worldwide have obesity, which is strongly linked to the risk of many cardiometabolic diseases, including coronary artery disease (CAD), stroke, and type 2 diabetes^1,2^. There are many biological mechanisms whereby obesity causes disease, including metabolic dysfunction, inflammation, and endothelial damage^3^. However, most of the factors mediating this relationship are not yet fully understood. Therefore, identifying modifiable mediators of this relationship could yield potential therapeutic targets, which may be targeted pharmaceutically or non-pharmaceutically, for example with lifestyle interventions. Circulating proteins are potential candidates because obesity strongly influences the level of plasma proteins^4,5^, and they play a critical role in disease development and progression. Moreover, circulating proteins can be measured and sometimes modulated^6^, and their levels can be used as a surrogate measure of target engagement in drug development programs. Therefore, understanding their role in disease could provide multiple avenues to lessen the impact of obesity on cardiometabolic disease.

One way to understand the role of circulating proteins in disease has been through observational epidemiology studies. However, such studies are not ideal for identifying causal mediators of disease because they are prone to bias from unmeasured confounders and reverse causation^7,8^, wherein the disease itself influences the protein level. What is therefore needed is a method to understand mechanisms of disease, while reducing such biases.

Mendelian randomization (MR) is a genetic epidemiology approach that can contribute to the understanding of the causal relationship between exposures and outcomes while minimizing the bias from confounding and avoiding reverse causation ^6-12^. MR can be described as a natural experiment somewhat analogous to randomized controlled trials (RCTs)^13^ because both rely upon randomization to reduce bias from confounding. In MR studies randomization is achieved through the random allocation of alleles at conception. Moreover, reverse causation can be theoretically avoided because genotype is always assigned prior to the onset of disease.

Despite these advantages, MR relies on three key assumptions^7,8^: there exist genetic variants that: (I) are associated with the risk factor of interest; (II) are not correlated with confounders of the exposure-outcome relationship; (III) affect the outcome only through the exposure (also known as lack of horizontal pleiotropy). Of these, the third assumption is the most problematic and can be a source of potential bias in MR. Nevertheless, when these main assumptions are met, MR can be a powerful tool to describe causal relationships in humans—free of model systems.

Advancements in large-scale proteomics have facilitated the discovery of genetic variants that influence plasma protein levels on a proteome-wide scale^14-16^. These genetic variants, referred to as protein quantitative trait loci (pQTLs), can be utilized in MR to estimate the causal effect of circulating protein levels on disease. Such methods have been successfully leveraged to prioritize therapeutic targets, including OAS1 for COVID-19^9,17^ and IL6R for both COVID-19^18,19^ and CAD^20^, and ANGPTL3 for CAD^21^. As drug discovery is costly and prone to failure^22^, proteo-genomics-based MR could play an important role since such studies could provide causal targets, which can be measured, thereby providing proximal read-out of drug target engagement, but also providing biomarkers for recruitment into clinical trials. Indeed, drugs with human genetics evidence are more likely to be successful in Phase II and III trials, and two-thirds of FDA-approved drugs in 2021 were supported by human genetics evidence^23,24^.

Furthermore, MR methods can be leveraged to understand mediators of the biological pathways connecting obesity with cardiometabolic disease when deployed in a two-step study design^25,26^. Step 1 begins by estimating the effect of BMI on protein mediators. Step 2 estimates the effect of the identified mediators on the outcome of interest (in this case, cardiometabolic diseases). Previously, we have successfully used this approach to identify a circulating protein, nephronectin, that mediates the impact of obesity on COVID-19 severity^27^.

In the present study, we conducted an integrative MR analysis of on a proteome-wide scale, screening 4,907 proteins, statistical colocalization, and mediation analysis to identify circulating proteins that mediate the effects of obesity on CAD, ischemic stroke, cardioembolic stroke, and type 2 diabetes. We then focused on collagen type VI α3 (COL6A3) as a potential target, performing multiple follow-up analyses, including replication and single-cell sequencing analysis. Additionally, we evaluated the actionability of COL6A3 by assessing the effect of reducing body fat on its circulating protein level in multivariable MR and also assessed the implication of reducing the identified proteins on a phenome-wide association study.

## Results

The overall study design and a summary of the results are illustrated in **Fig. 1**. The study consisted of four main sections:

**Figure 1.**
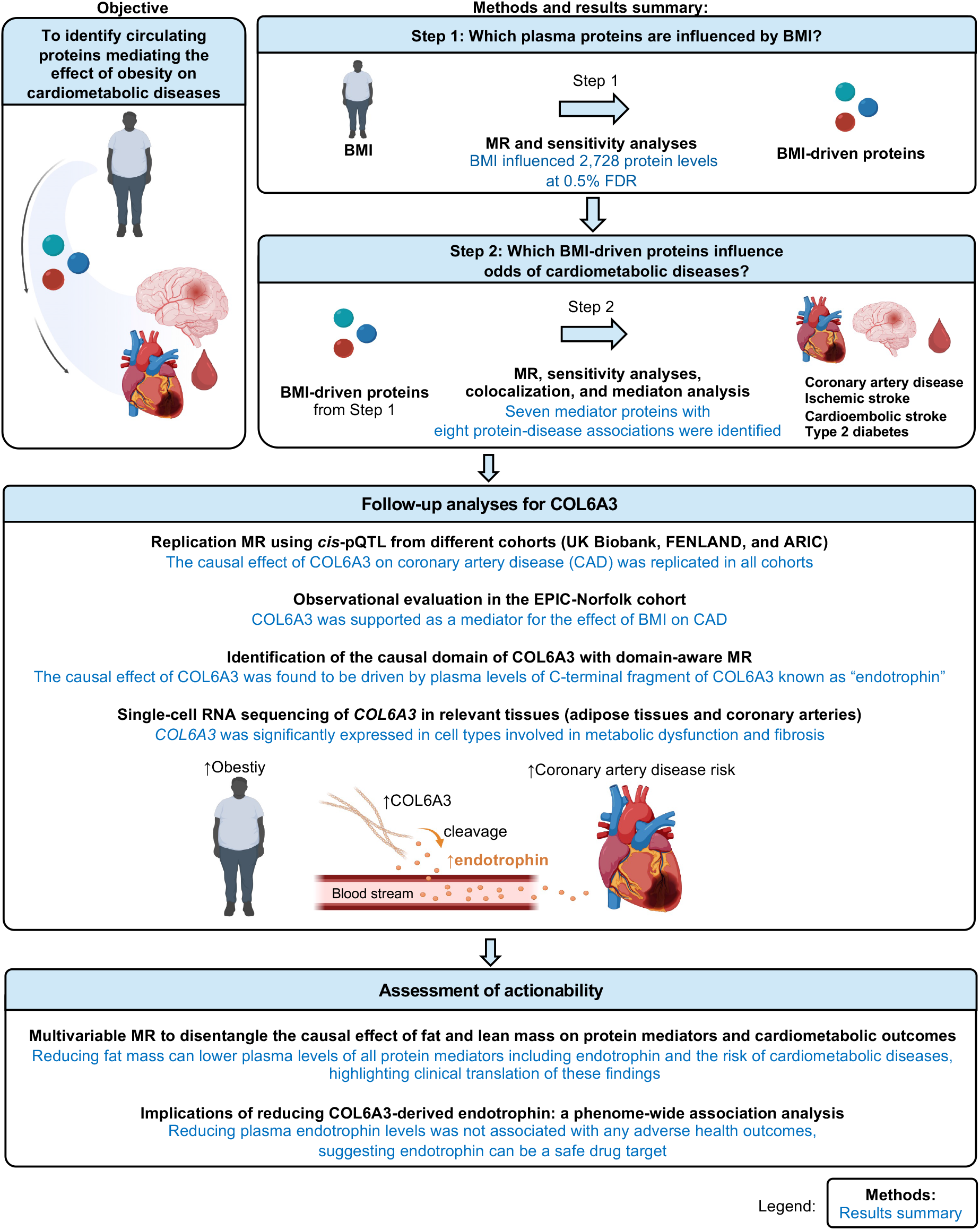
Study design. To identify proteins that mediates the effect of obesity on cardiometabolic diseases, we used a two-step approach. In Step 1 Mendelian randomization (MR), we assessed the effect of body mass index (BMI) on 4,907 plasma proteins, which led to the identification of 2,714 proteins influenced by BMI (referred to as “BMI-driven proteins”) using two-sample MR. In Step 2 MR, we assessed the effect of these BMI-driven proteins on cardiometabolic diseases, again using two-sample MR. In the subsequent sections, we conducted follow-up analyses of COL6A3 and evaluated the potential for actionability of this protein and other mediators we identified. BMI: body mass index, *cis*-pQTL: *cis*-acting quantitative trait loci.

1. Step 1 MR, which evaluated the causal effect of body mass index (BMI) on the levels of circulating plasma proteins. We also evaluated the consistency of MR findings when BMI and body fat percentage were used as the exposures.
2. Step 2 MR, which assessed the causal effects of BMI-driven proteins on four cardiometabolic outcomes (CAD, ischemic stroke, cardioembolic stroke, and type 2 diabetes).
3. Follow-up analyses for COL6A3 and its cleavage product, known as endotrophin, which assessed its role in CAD.
4. Assessment of clinical actionability for COL6A3-derived endotrophin and other protein mediators by reducing body fat mass.

Each of these four steps and their results is described in detail below.

### 1) Step 1 MR: Identification of the causal effect of BMI on plasma protein levels

We evaluated the causal effect of BMI on 4,907 circulating proteins using the SomaScan v4 aptamer binding assay (SomaLogic, Boulder, CO). For clarity, we will refer to protein-targeting aptamers as “proteins” unless otherwise specified. We performed causal inference using two-sample MR, to estimate the effect of an exposure on an outcome of interest using two separate genome-wide association studies (GWAS); one for the BMI and the second for circulating proteins^13^ (**Methods**). Specifically, we used the GWAS of BMI from the GIANT and UK Biobank consortia^28^ (*n* = 681,275 individuals) and circulating protein levels from the deCODE study^15^ (*n* = 35,559 individuals). In both studies we included only participants of European genetic ancestry (**Supplementary Table 1**). We performed two-sample MR, using the inverse variance weighted method as the primary analysis and then filtered these results dependent upon sensitivity analyses, including tests for heterogeneity, directional horizontal pleiotropy, and reverse causation. We used false discovery rate (FDR) correction with 0.5% as a strigent threshold for significance, given that many protein levels are correlated with each other and therefore a Bonferroni correction would be overly conservative (see **Methods**). No evidence of weak instrumental variables (suspected when F-statistics < 10) were found (**Supplementary Table 2**).

We found that BMI influenced 2,728 proteins, passing tests of significance, heterogeneity, and directional pleiotropy (**Supplementary Table 3**). However, among them, 14 showed evidence of reverse causation, wherein the protein influenced BMI (**Supplementary Table 4**), and these 14 proteins were removed from further analyses. Thus, we identified a total of 2,714 plasma proteins that are influenced by BMI. Hereafter, these 2,714 proteins are referred to as BMI-driven proteins (**Fig. 2a, 2b, and 2c**).

**Figure 2.**
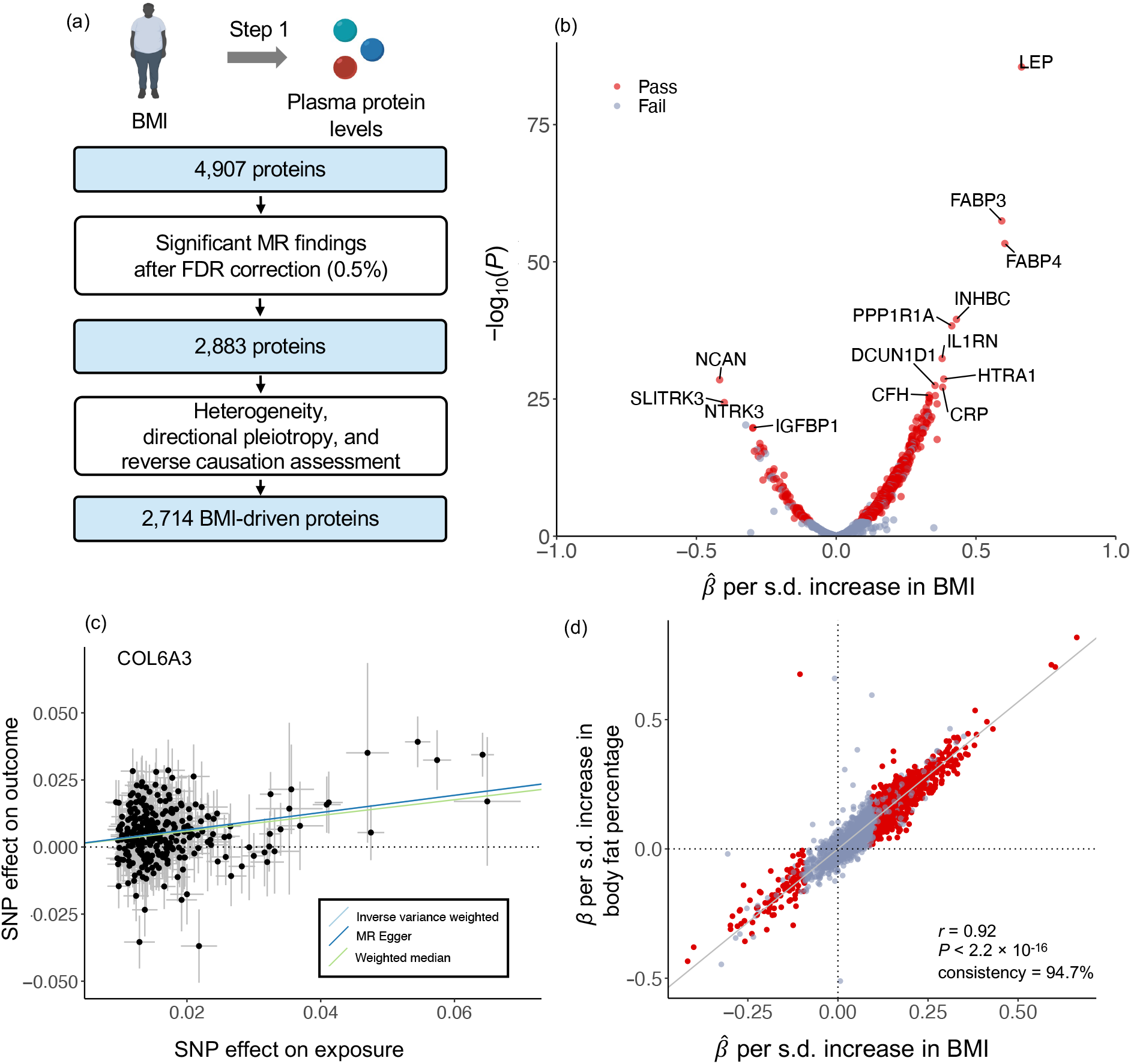
MR analyses for the effect of BMI on plasma protein levels. (a) Flow diagram outlines Step 1 Mendelian randomization (MR). (b) A volcano plot illustrates the effect of BMI on each plasma protein from MR analyses using the inverse variance weighted method. The x-axis represents beta estimates, and the y-axis represents -log_10_(*P*) values from MR results. Red dots represent proteins that passed all tests, including significance with a false discovery rate (FDR) < 0.5%, as well as tests for heterogeneity, directional pleiotropy, and reverse causation. Grey dots represent proteins that failed any of these tests. (c) MR scatter plot shows the effect of BMI on plasma levels of COL6A3 using the inverse-variance weighted method (primary analysis), weighted median, or MR-Egger slope methods. (d) Directional consistency between MR results for the effect of BMI on plasma proteins and MR results for the effect of body fat percentage on plasma protein levels using the inverse variance weighted method. The x-axis denotes beta estimates from MR results, and *r* denotes Pearson’s correlation.

Additionally, we performed MR to evaluate the effect of body fat percentage on the same 4,907 plasma proteins (**Methods**). We did this because body fat percentage is considered to be a more direct proxy of obesity, whereas BMI is an easy-to-measure, clinically relevant proxy.^29^ However, the sample size available to assess the genetic determinants of BMI is larger than that of body fat percentage, provide more precise estimates. We found that body fat percentage influenced 94.7% of all BMI-driven proteins with the same direction of effect as BMI **(Fig. 2d**), illustrating a high concordance of results between the two different measures of obesity (*r* = 0.93; *P* < 2.2 × 10^-16^). Given the high concordance between MR results from BMI and body fat percentage, we proceed to Step 2 MR with BMI-driven protein results.

### 2) Step 2 MR: Identification of the causal effect of BMI-driven proteins on cardiometabolic diseases

Next, we estimated the causal effect of these BMI-driven proteins on CAD, ischemic stroke, cardioembolic stroke, and type 2 diabetes, again using two-sample MR (**Fig. 3a**). We used the BMI-driven protein levels identified in Step 1 MR as exposures. The outcomes were CAD, ischemic stroke, cardioembolic stroke, and type 2 diabetes (see **Methods**). To minimize the risk of bias from horizontal pleiotropy, we used *cis*-acting protein quantitative trait loci (*cis*-pQTLs) identified from 35,559 individuals from the deCODE study^15^ as instrumental variables. In this context, instrumental variables are genetic variants that influence the exposure (i.e., circulating protein levels). We have defined *cis*-pQTLs as pQTLs that reside within a ± 1 Mb region around a transcription start site of a protein-coding gene. Since such *cis*-pQTLs would be likely to directly influence the circulating protein level by influencing the transcription or translation of mRNA from the gene that encodes the protein, they are less prone to bias from horizontal pleiotropy. Horizontal pleiotropy produces bias from the genetic variant influences the outcome independently of the circulating protein level.

**Figure 3.**
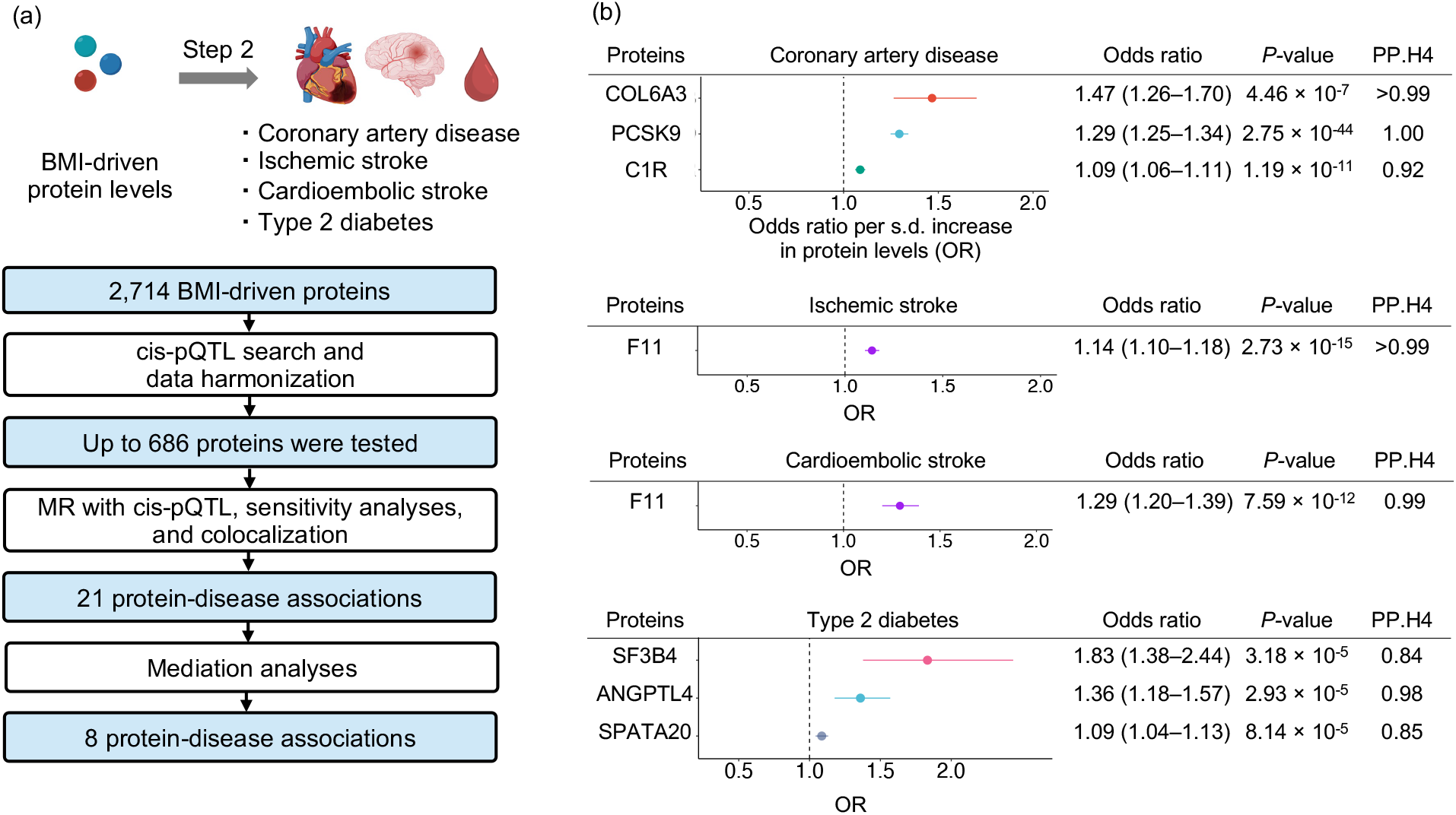
MR analyses for the effect of BMI-driven proteins on cardiometabolic diseases. (a) Flow diagram of the Step 2 Mendelian randomization (MR) analyses. (b) Forest plots for the effect of body mass index (BMI)-driven proteins on four cardiometabolic diseases (coronary artery disease, ischemic stroke, cardioembolic stroke, type 2 diabetes). The MR analyses were conducted using the largest available GWAS of coronary artery disease^30^ (181,522 cases and 1,165,690 controls), ischemic stroke (34,217 cases and 2,703,029 controls), cardioembolic stroke^31^ (7,193 cases and 2,703,029 controls), and type 2 diabetes^32^ (80,154 cases and 853,816 controls).

To further reduce the risk of horizontal pleiotropy, we restricted instrumental variables to genetic variants that were *cis*-pQTLs to only one protein. To do so, we removed variants associated with more than two proteins in a *cis*-acting manner (**Fig. 3a; see Methods**). For the outcomes, we used the largest available GWAS for CAD^30^ (181,522 cases and 1,165,690 controls), ischemic stroke, and cardioembolic stroke^31^ (34,217 ischemic stroke cases, 7,193 cardioembolic stroke cases, and up to 2,703,029 controls), and type 2 diabetes^32^ (80,154 cases and 853,816 controls).

Following MR with *cis*-pQTLs and sensitivity analyses (heterogeneity, pleiotropy, and reverse causation assessment), we performed colocalization to evaluate whether the pQTL of the protein of interest and the disease outcome shared a single causal variant around a 1-Mb (± 500 kb) region surrounding the lead *cis*-pQTL. As different linkage disequilibrium (LD) structures across different study populations may lead to bias in the MR estimates, the presence of a shared single causal variant between the pQTL and the disease outcome can increase the robustness of MR findings (see **Methods**).

After MR with *cis*-pQTLs, sensitivity analyses, and colocalization, we identified 21 protein-disease associations that passed both step 1 and step 2 MR (**Supplementary Table 5**), including collagen type VI α3 (COL6A3) and PCSK9 for CAD, F11 for ischemic and cardioembolic stroke, and SF3B4 for type 2 diabetes. Among these proteins, COL6A3 was associated with the highest odds of CAD per one standard deviation (s.d.) increase in the protein levels (odds ratio (OR) = 1.47, 95% CI: 1.26–1.70, *P* = 4.7 × 10^-7^). We note that the finding of PCSK9 serves as a “positive control” and illustrates the utility of this method as PCSK9 is a well-known drug target, and its inhibition has been shown to reduced cardiovascular outcomes in multiple clinical trials^33-35^. Full results for CAD, ischemic stroke, cardioembolic stroke, and type 2 diabetes are provided in **Supplementary Table 6–9.**

As an additional filtering step, we performed mediation analyses for the identified protein-disease associations. To do this, we used the product of coefficients method^27,36-38^ (**Methods**). Given that BMI increases the risk of cardiometabolic diseases (*β*_BMI-to-cardiometabolic_ > 0 in **Extended Fig. 1**; **Supplementary Table 10)**, we restricted the analysis to proteins that increased the risk of cardiometabolic diseases through their mediation pathway (*β*_BMI-to-protein_ × *β*_protein-to-cardiometabolic_> 0 in **Extended Fig. 1; Supplementary Fig. 10**). Among the 21 protein-disease associations, 8 met this condition. Notably, all eight protein-disease associations were supported by mediation analyses, suggesting that the effect of BMI on the cardiometabolic outcome was mediated, at least partially, by the circulating protein (**Figure 3b; Supplementary Table 10**).

### Follow-up analyses of COL6A3 (collagen type VI α3)

Circulating COL6A3 levels had the strongest effects on CAD across all the mediators of the relationship between BMI and this outcome. We therefore sought to further test the hypothesis that COL6A3 mediates the relationship between obesity and cardiometabolic disease using analyses from orthogonal resources.

#### Replication MR using cis-pQTL from different cohorts

We evaluated whether the causal relationship between COL6A3 and CAD could be replicated using different sources of *cis*-pQTLs from other cohorts. For this, we conducted two-sample MR using *cis*-pQTLs from three additional cohorts: UK Biobank^39^ (*n* = 35,571 individuals), Fenland^14^ (*n* = 10,708 individuals), and ARIC^16^ (*n* = 7,213 individuals). MR in all cohorts supported the causal effect of COL6A3 levels on CAD, in the same direction **(Supplementary Table 11**). Specifically, each s.d. increase in COL6A3 was associated with increased odds of CAD in UK Biobank^39^ (OR = 1.30, 95% CI: 1.17–1.45, *P =* 2.4 × 10^-6^), Fenland (OR = 1.23, 95%CI: 1.12–1.35, *P =* 8.9 × 10^-6^), and ARIC (OR = 1.09, 95%CI: 1.05–1.13, *P =* 1.6 × 10^-5^). Notably, UK Biobank used Olink Explore 3072 assay^39^, whereas deCODE^15^, Fenland^14^, and ARIC^16^ used SomaScan v4 assay. Hence, concordant MR results using *cis*-pQTLs from the different studies from two different proteomic platforms further strengthened the evidence that COL6A3 partially mediates the relationship between obesity and CAD.

#### Observational epidemiological evaluation in the EPIC-Norfolk cohort

If testing a hypothesis using different designs yields similar results, it is less likely that the results are due to bias specific to one of the study designs. This is because different study designs have different bias architectures and concordant results across study designs strengthens causal inference because it is less likely that a single source of bias generated the results. Such testing has been referred to as a triangulation of evidence. We therefore performed observational association analysis with a randomly selected sub-cohort of the EPIC-Norfolk study (*n* = 872), which included 207 prevalent or incident cases of CAD (see **Methods**). EPIC-Norfolk is a population-based cohort from the United Kingdom. We found that increased BMI was associated with increased plasma levels of COL6A3 (*β* = 0.06, 95% CI: 0.04–0.08, *P =* 8.5 × 10^-12^), and a s.d. increase in plasma COL6A3 levels was associated with increased odds of CAD (OR = 1.34, 95% CI: 1.12–1.59, *P =* 1.1 × 10^-3^). The mediation analysis supported that plasma COL6A3 levels partially mediated the effect of BMI on CAD (**Supplementary Table 12**).

Given the robustness of these findings, we then explored the potential mechanism whereby COL6A3 may influence CAD.

#### Identification of the causal domain of COL6A3

Cleavage of proteins can influence their biological mechanism^40^. Previous studies have shown that the C-terminal domain, also known the Kunitz domain, of COL6A3 is proteolytically cleaved to form a biologically active fragment known as “endotrophin”. Endotrophin is produced in multiple tissues, including adipose tissue^40,41^. Endotrophin strongly induces fibrosis and inflammation, and recent evidence suggests that it is involved in obesity-induced metabolic dysfunction^40-45^ (**Fig 4a**). Therefore, we evaluated whether this particular domain of COL6A3 is driving its effect on CAD.

**Figure 4.**
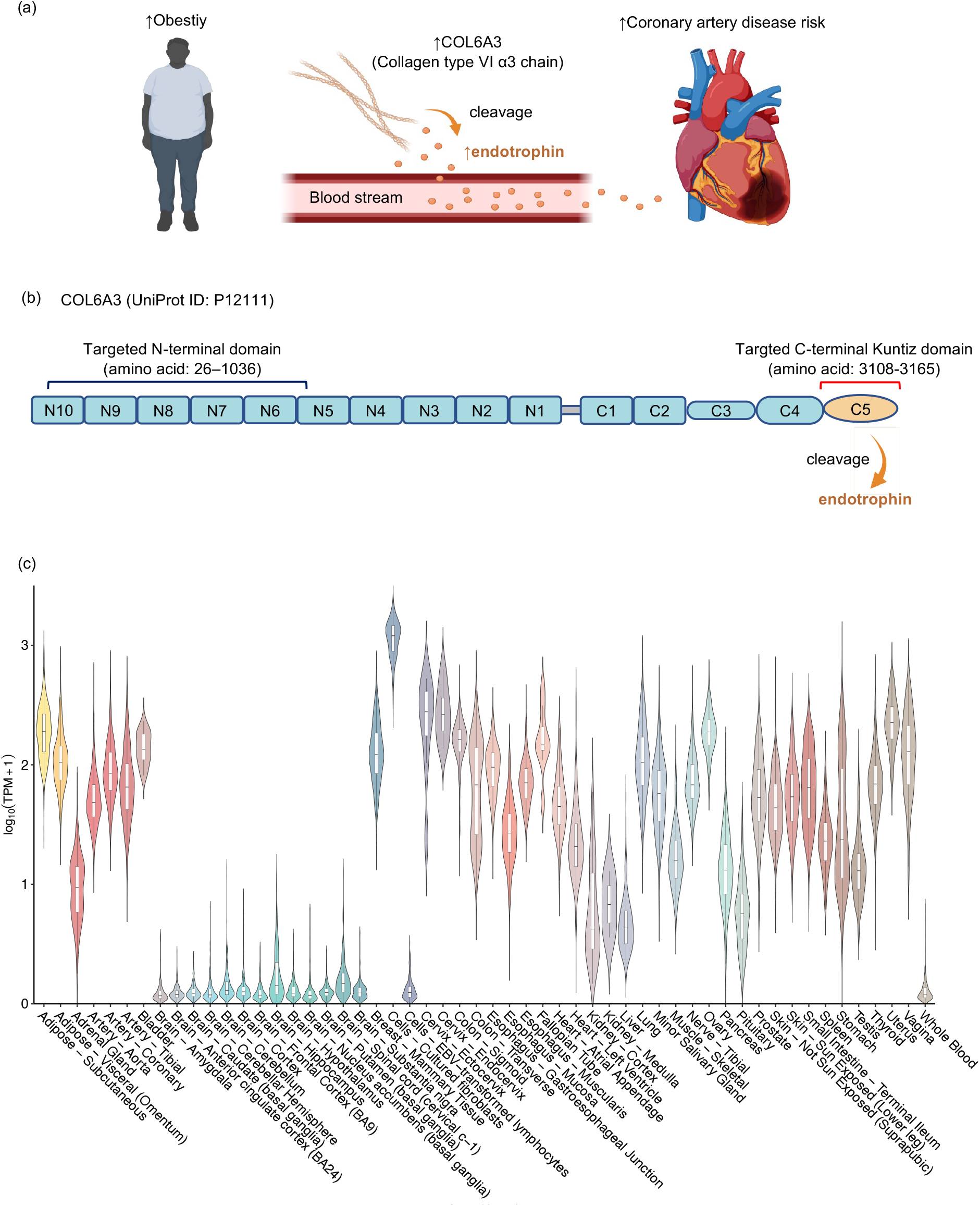
Follow-up analyses for collagen type VI α3 (COL6A3). (a) Schematic illustration of proposed relationship between obesity, COL6A3 (Collagen type VI α3 chain), endotrophin, and coronary artery disease. Obesity leads to increased production of COL6A3, whose C-terminal is cleaved into an active form termed endotrophin, which increases the risk of coronary artery disease. (b) Schematic diagram of COL6A3 (UniProt ID: P12111). COLA3 consists of a short collagenous region flanked by multiple von Willebrand factor type A (vWF-A) modules (N1–N10 in the N-terminal and C1,2 in the C-terminal). There are three additional C-terminal domains unique to COL6A3 (C3–C5), which are not present in other collagen type VI families. The most C-terminal domain (C5) is cleaved into a soluble protein termed endotrophin. The two amino acid sequences targeted by the aptamers are as follows: the N-terminal-binding aptamer targets the amino acid sequence 26–1036 (uncleaved section), while the C-terminal aptamer targets the amino acid sequence 3108–3165 (cleaved section). The figure has been modified from ref^81,82^. (c) *COL6A3* expression profile in human tissues in GTEx v8^48^. *COL6A3* expression levels were represented on a log transcript per 10 thousand plus one (TPM + 1) scale.

The SomaScan v4 assay measures target protein levels using aptamers, which are short, single-stranded DNA or RNA molecules that can selectively bind to the target protein^46^. SomaScan v4 assay has two separate aptamers targeting two domains of COL6A3, the N-terminal and C-terminal (Kunitz domain) (**Methods).** These two separate aptamers thus allowed us to disentangle the effects of the N-terminal and C-terminal containing fragments of COL6A3.

Intriguingly, we found that the aptamer binding the C-terminal of COL6A3 (**Fig. 4b)** was associated with an increased risk of CAD (OR = 1.46 per s.d. increase in the protein level, 95% CI: 1.37–1.93, *P* = 2.7 × 10^-8^), whereas the aptamer binding the N-terminal (i.e., the non-cleaved portion of COL6A3) was not associated with the risk of CAD (OR = 1.06, 95% CI: 0.96–1.18, *P* = 0.22) in domain-aware MR (**Supplementary Table 13**). These findings suggest that the C-terminal of COL6A3, which is cleaved into endotrophin, explains the effect of COL6A3 on CAD and the aptamer binding to the C-terminal of COL6A3 may be capturing the plasma levels of endotrophin or endotrophin-containing fragments. In the remainder of the manuscript, we refer to such fragments as endotrophin for clarity.

To further test the hypothesis that endotrophin is responsible for COL6A3’s effect upon CAD, we tested whether *cis*-pQTLs from the Olink Explore 3072 assay^39,47^ for COL6A3 were associated with CAD. The Olink Explore 3072 assay uses a polyclonal antibody to target the C-terminal (Kuniz domain) of COL6A3. The *cis*-pQTL (rs1050785) from UK-Biobank, which uses the Olink platform, was in high linkage disequilibrium (*R^2^* = 0.73) with *cis*-pQTL (rs11677932) of the C-terminal-targeting aptamer from the deCODE study but not in LD (*R^2^* = 0.0) with the *cis*-pQTL of the N-terminal-targeting aptamer of COL6A3 (rs2646260). We found that the *cis*-pQTL from the Olink platform was strongly associated with increased odds of CAD (OR = 1.32, 95%CI: 1.16– 1.50, *P =* 1.75 × 10^-5^) (**Supplementary Table 13**), which was consistent with the finding using SomaScan v4 assay’s aptamer binding the C-terminal of COL6A3. Taken together, these results provide evidence from orthogonal proteomic assays that circulating levels of C-terminus COL6A3-derived endotrophin likely explain the effect of COL6A3 levels on CAD.

Moreover, domain-aware MR analysis revealed that the aptamer targeting the C-terminal of COL6A3 (cleaved portion) was more strongly increased by an increase in BMI (*β* = 0.32, 95% CI: 0.26–0.38, *P =* 3.7 × 10^-24^) than the aptamer targeting N-terminal (uncleaved portion) (*β* = 0.10, 95% CI: 0.04–0.16, *P =* 2.1 × 10^-3^), as shown by non-overlapping confidence intervals. These findings indicate that an increase in BMI could increase both the expression of *COL6A3* and its cleavage, but has a preferential effect on the cleavage of COL6A3 into endotrophin.

#### COL6A3 expression analyses

We next explored the tissues in which COL6A3 is expressed using GTEx v8, which is a compendium of expression data from 49 tissues across 838 individuals^48^. In GTEx v8 (https://gtexportal.org/), *COL6A3* was significantly expressed in multiple tissues, including adipose tissue and coronary arteries when compared to the whole blood (*P* < 0.001) (**Fig. 4c**). Therefore, it is possible that these tissues may locally produce COL6A3 and consequently its cleavage product, endotrophin. While tissue-level examination of expression is helpful, such methods do not permit resolution to the cellular level. Considering that the adipose tissue is reported to be the primary source of COL6A3^45^ and that the coronary artery is the location of primary lesions in CAD^49^, to better understand the cell type of origin of COL6A3 we analyzed single-cell *COL6A3* expression in human white adipose tissues^50^ (SCP1376 at https://singlecell.broadinstitute.org/) and coronary arteries in patients with CAD^49^ (GSE131780 at https://www.ncbi.nlm.nih.gov/geo/).

In single-cell sequencing, *COL6A3* was significantly enriched in adipose progenitor/stem cells of adipose tissues when compared to other cell types in adipose tissues (permutation *P* < 0.001; see **Methods**) (**Fig. 5a)**. Given that these cell populations play critical roles in maintaining adipose tissue and metabolic function^51,52^, the findings indicate that metabolic dysfunction may be an underlying biological mechanism whereby COL6A3 influences CAD. Additionally, we found that COL6A3 was significantly expressed in fibroblasts, which plays a key role in the atherosclerosis of the coronary artery^53^, when compared to other cell types in the coronary artery (permutation *P* < 0.001; see **Methods**) (**Fig. 5b)**. Taken together, these findings suggested that these cell types may be responsible for the local production of COL6A3 in these tissues.

**Figure 5.**
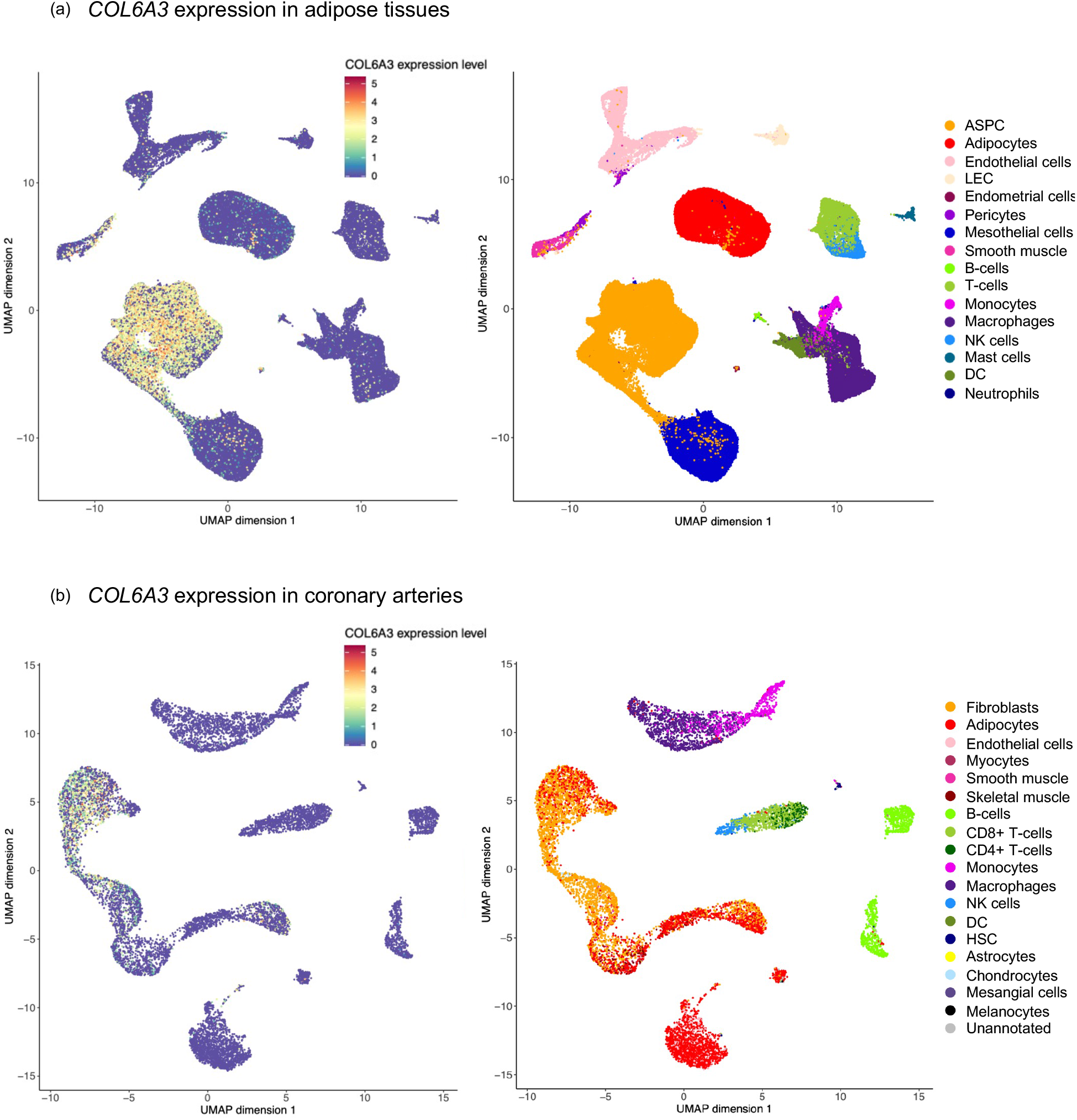
Single-cell sequencing analyses of *COL6A3*. *COL6A3* expression patterns in the adipose tissues (a) and coronary arteries (b). We obtained single-cell transcriptomic data of human adipose tissue from Emont et al.^50^ (SCP1376 at https://singlecell.broadinstitute.org/) and the data of coronary arteries from Wirka et al.^49^ (GSE131780 at the Gene Expression Omnibus database https://www.ncbi.nlm.nih.gov/geo/). ASPC: adipose stem and progenitor cells, LEC: lymphatic endothelial cells, NK: natural killer cells, DC: dendritic cells.

### Assessment of clinical actionability

While identifying mediators of the effect of obesity on cardio-metabolic disease is relevant, such targets could become clinically relevant if their modification through weight loss or other methods influenced disease outcomes. We therefore explored whether reducing fat mass and/or increasing lean mass could improve plasma COL6A3-derived endotrophin and other protein levels, thereby reducing the risk of cardiometabolic diseases. For this, we used multivariable MR to evaluate the independent effects of body fat and lean mass (i.e., body fat-free mass) on the protein mediators and cardiometabolic disease outcomes (**Methods**).

We found that an s.d. increase in fat mass was independently associated with increased plasma levels of all protein mediators (COL6A3-derived endotrophin, F11, PCSK9, C1R, SPATA20, SF3B4, and ANGPTL4) (**Fig. 6b and Supplementary Table 14**) and increased odds of type 2 diabetes, CAD, and ischemic stroke. On the contrary, an s.d. increase in lean mass was independently associated with decreased plasma levels of some protein mediators including F11 and PCSK9 (**Fig. 6b and Supplementary Table 15**).

**Figure 6.**
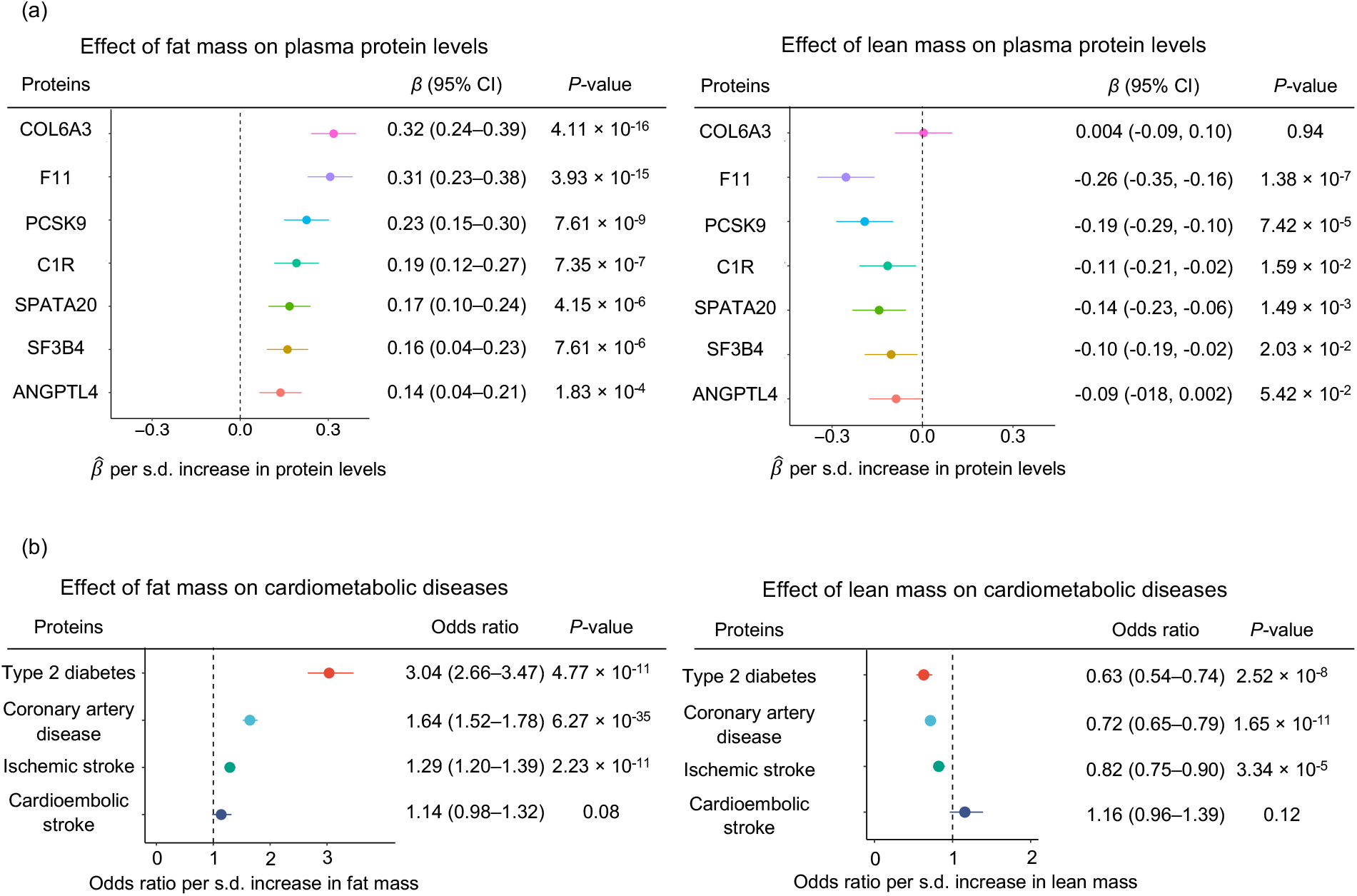
Multivariable MR analysis for evaluating the independent effects of fat mass and lean mass on plasma NPNT levels (a) and cardiometabolic diseases (b). We performed multivariable Mendelian randomization (MR) using fat mass and lean mass as exposures and plasma protein levels of the seven protein mediators or cardiometabolic diseases as outcomes.

This has important clinical implications for actionability because interventions such as exercise, appropriate diet, or weight loss drugs such as the GLP-1 receptor agonist semaglutide and GLP-1/GIP co-agonist tirzepatide, which reduces body fat mass more than lean mass^54,55^, could be effective in improving these protein levels and subsequently decreasing the risk of cardiometabolic diseases. However, future clinical trials are needed to confirm this hypothesis.

Lastly, we evaluated whether reducing COL6A3-derived endotrophin is associated with any adverse health outcomes using a phenome-wide association analysis in the UK Biobank, FinnGen, and the GWAS catalog. We did this because clinical trials for some drug candidates have been terminated due to unexpected adverse events in later stages of the trials^22,56^; thus, understanding the potential effects of perturbing the target on a phenome-wide level may to anticipate possible adverse events. Therefore, we assessed whether reducing COL6A3-derived endotrophin levels may have any implications on other traits. For this, we queried traits associated with the lead *cis*-pQTL of COL6A3 (rs11677932) from the deCODE study (a proxy for the COL6A3’s C-terminal-derived endotrophin) in data from UK Biobank, FinnGen, and GWAS catalog using the Open Target Genetics (https://genetics.opentargets.org/) at *P* < 1.0 × 10^-5^. The phenome-wide association analysis revealed that decreased plasma levels of COL6A3-derived endotrophin (A-allele of rs11677932; *β* = -0.07, *P* = 1.5 × 10^-14^) was associated with decreased risk of coronary atherosclerosis (*β* = -0.05, *P* = 1.0 × 10^-5^), increased heel bone mineral density (*β* = 0.02, *P* = 2.9

× 10^-12^), and increased lung function (FEV1/FVC) (*β* = 0.02, *P* = 5.2 × 10^-13^) in addition to reduced risk of CAD (*β* = -0.03, *P* = 2.9 × 10^-12^) (**Supplementary Table 16**).This suggests that decreasing COL6A3-derived endotrophin may decrease the risk of multiple morbidities, including coronary atherosclerosis, CAD, bone mineral density, and lung function, offering COL6A3-derived endotrophin as an attractive therapeutic target.

## Discussion

Obesity is a major risk factor of multiple diseases, and therapies are required that reduce its clinical consequences. Here, we identified seven protein mediators (from eight protein-disease associations) that partially mediate the effect of obesity on cardiometabolic diseases in humans. All of these protein levels, including COL6A3, could potentially be improved through body fat reduction, illustrating their possible clinical actionability. Furthermore, triangulation of evidence with multiple follow-up analyses indicated that endotrophin, which is derived from the cleavage of COL6A3, drives a part of the effect of obesity on CAD. These findings provide insights into how obesity causes cardiometabolic disease and provide circulating proteins that could be investigated as potential drug targets to lessen the public health burden of obesity.

The major finding of this study is the mediating role of endotrophin in the effect of obesity on CAD in humans. Previous studies reported endotrophin as an important hormone that induces metabolic dysfunction, fibrosis, and inflammation in rodent models^40-42,57^, and cross-sectional studies in humans have found that increased circulating endotrophin level was observationally associated with cardiovascular events and all-cause mortality^43-45,58^. However, cross-sectional observational studies cannot disentangle cause and consequence. Therefore, our study, which utilized MR to make causal inferences, provides evidence that endotrophin acts as a causal mediator for the relationship between obesity and CAD in humans. Considering our findings that reducing COL6A3 and its cleaved product, endotrophin, can reduce the risk of CAD without apparent adverse health outcomes, directly targeting endotrophin can be an attractive therapeutic approach, and it may be particularly effective in individuals with obesity.

Notably, we found that the aptamer targeting C-terminal of COL6A3 (also called the Kunitz domain, which is cleaved into endotrophin) was more strongly affected by an increase in BMI than the aptamer targeting the N-terminal. This indicates that obesity may increase both *COL6A3* expression and the cleavage of COL6A3, but with a preferential influence on the cleavage of COL6A3 into endotrophin, leading to an increase in endotrophin levels. Several studies using mice models have shown that the bone morphogenetic protein 1 (BMP1)^41^, matrix metallopeptidase 14 (MMP14)^59^, and other MMPs^60^ can release the C-terminal of COL6A3 as endotrophin after proteolytic cleavage. However, as these studies were conducted using rodent models, further research is needed to establish whether the same applies to humans. Despite this, inhibition of BMP1 reduces scar formation and supports the survival of cardiomyocytes^61^, which may be partly due to lower levels of endotrophin. Nevertheless, BMP1 also cleaves other procollagens into mature collagens, which introduces pleiotropy. Therefore, more research is necessary to determine how to selectively inhibit the cleavage of the C-terminal of COL6A3 to reduce endotrophin levels.

Our study also illuminated other proteins, such as ANGPTL4, which mediate the relationship between obesity and type 2 diabetes. Previous studies have shown that ANGPTL4 inhibits lipoprotein lipase^62^, thereby reducing triglyceride levels^63^. Additionally, ANGPTL4 has also been implicated as an important player in obesity-induced glucose intolerance^62-66^, consistent with our findings. Currently, an ANGPTL4 inhibitor, which is hepatocyte-targeting GalNAc-conjugated antisense oligonucleotides that downregulate ANGPTL4 levels in liver and adipose tissue, is in phase 1 clinical trial for hypertriglycedemia^67^. Our research indicates that this drug may be tested for the prevention of type 2 diabetes, and further clinical trials are required to evaluate the safety and efficacy of ANGPTL4 inhibition in humans. Another notable finding is F11 (coagulation factor XI) as a mediator of the effect of obesity on cardiometabolic disease. F11 is a critical player in the coagulation pathway and has been identified as causal for stroke by multiple studies^6,68^. However, few studies highlighted its role as a mediator. Currently, the F11 inhibitor, abelacimab^69^, is in phase III clinical trial for venous thromboembolism (NCT05171049 at https://www.clinicaltrials.gov/). Our findings suggest that this drug may be effective for reducing the risk of ischemic stroke, especially for individuals with obesity.

This study has important limitations. First, we focused on analyzing data solely from European-ancestry individuals to prevent confounding by population stratification. While the ARIC cohort reported *cis*-pQTL for individuals of African ancestry^16^, the sample size (*n* = 1,871) is still limited when compared to data for those of European ancestry (deCODE study; *n* = 35,559). The same applies to CAD GWAS, with 181,522 CAD cases in European ancestry individuals^70^ compared to only 17,247 cases in African ancestry individuals^71^. This limited sample size in African ancestry individuals reduces the statistical power of MR analysis. Therefore, further efforts are needed to increase the sample size of non-European-ancestry pQTL data. Second, we did not perform sex-stratified analysis due to the unavailability of sex-specific datasets. Third, while the mediation analyses results with both MR and observational evaluation in EPIC-Norfolk provided additional evidence supporting COL6A3-derived endotrophin as a causal mediator, it should be noted that the mediation analyses are based on additional assumptions^72^. Therefore, we used them as one of several orthogonal validation methods. Fourth, we did not explore the molecular mechanism whereby these proteins mediated the effect. Finally, although we triangulated multiple lines of evidence to propose several promising therapeutic targets that mediate an important proportion of the effect of obesity on cardiometabolic diseases (e.g., COL6A-derived endotrophin and ANGPTL4), future clinical trials are required to explore the effect of pharmacologically influencing these protein levels.

## Conclusions

These results provide actionable insights into how circulating proteins mediate the effect of obesity on cardiometabolic diseases. Our study highlights the importance of body fat reduction to reduce the risk of cardiometabolic diseases and offers potential therapeutic targets, including COL6A3-derived endotrophin, which may be prioritized for drug development.

## Methods

### Step 1 MR

#### MR to evaluate the effect of BMI on plasma protein levels

We performed two-sample MR using BMI as exposure and circulating protein levels as outcomes. The BMI exposure data came from a meta-analysis GWAS of UK Biobank and GIANT involving 693,529 European-ancestry individuals^28^ (**Supplementary Table 1**). For the outcomes, we used a GWAS of protein levels from the deCODE study^15^, measuring 4,907 proteins in 35,559 individuals of European ancestry using the SomaScan assay v4 from SomaLogic (Boulder, Colorado, USA).

We performed two-sample MR using genome-wide significant and independent single nucleotide polymorphisms (SNPs) with *P* < 5 × 10^-8^ and *r^2^* < 0.001 as instrumental variables. We excluded SNPs in the human major histocompatibility complex region because of their complex linkage disequilibrium structures. Clumping was performed using PLINK v1.9 (https://www.cog-genomics.org/plink/) with 10-Mb window. When the instrumental variable SNPs were not present in the outcome GWAS, we identified proxy SNPs with *r^2^* ≥ 0.8 using snappy v1.0 (https://gitlab.com/richards-lab/vince.forgetta/snappy/). To reduce the risk of weak instrument bias, we calculated F-statistics and evaluated whether they were above ten^73,74^ (**Supplementary Table 2)**.

After harmonizing the exposure and outcome GWAS, we performed two-sample MR analysis using the inverse variance weighted method with a random-effects model as the primary analysis, implemented using TwoSampleMR v0.5.6. We set FDR < 0.005 (0.5%) as a stringent threshold for significance. We used FDR correction, given that many proteins are correlated with each other and that a Bonferroni correction can be overly conservative in such situations. However, we used a strict threshold of 0.5% instead of a conventional threshold of 5% to reduce false positive findings, as our intention was not to generate a complete list of potential associations, but rather to generate a smaller set of high-confidence findings. We used weighted median, weighted mode, and MR-Egger slope as supplementary analyses to evaluate the directional concordance of the effect. Heterogeneity was tested using the *I^2^* statistic with results of *I*^2^ > 50% and heterogeneity *P* < 0.05 considered as substantial heterogeneity. Directional horizontal pleiotropy was tested using the MR-Egger intercept test, and results with *P* < 0.05 were considered to indicate the presence of directional horizontal pleiotropy.

For reverse MR, wherein we examined the effect of plasma protein levels on BMI, we performed two-sample MR using *cis*-pQTLs variants from the deCODE study as exposures and BMI GWAS from UK Biobank as an outcome. We used the inverse variance weighted method or the Wald ratio method when only one SNP was available. We used FDR < 0.5% as a threshold for significance. For BMI GWAS, we used data from the UK Biobank instead of the meta-analysis GWAS of UK Biobank and GIANT because a number of *cis*-pQTL SNPs were not available in the latter due to the stringent quality control process of the meta-analysis.

#### MR to evaluate the effect of body fat percentage on plasma protein levels

While BMI is an easily measurable, clinically relevant proxy of obesity with the largest GWAS, body fat percentage is considered a more direct measurement of body fat accumulation. Thus, a high concordance between the BMI and body fat accumulation MR results may strengthen the inference from the findings of Step 1 MR for BMI.

Therefore, we performed two-sample MR using body fat percentage as exposure and plasma protein levels as outcomes. We used GWAS of body fat percentage in 454,633 European-ancestry individuals from UK Biobank (Accession ID: ukb-b-8909 at IEU OpenGWAS project) and the same protein levels for GWAS from the deCODE study as used in Step 1 MR.

### Step 2 MR

#### MR with cis-pQTL to evaluate the effect of BMI-driven proteins on disease outcomes

Next, we performed two-sample MR using circulating protein levels as exposures and cardiometabolic diseases as outcomes, separately for each disease outcome. We used *cis*-pQTL variants from the deCODE study in 35,559 European-ancestry individuals^15^ as the instrumental variables. The *cis*-pQTL was defined as pQTL located within 1 Mb (± 1Mb) from the transcription start site of the corresponding protein-coding gene. For the outcome, we used the largest available GWAS of CAD^30^ (181,522 CAD cases and 1,165,690 controls), ischemic stroke, and cardioembolic stroke^31^ (34,217 ischemic stroke cases, 7,193 cardioembolic stroke cases, and up to 2,703,029 controls), and type 2 diabetes^32^ (80,154 type 2 diabetes cases and 853,816 controls). After data harmonization, we estimated the effect of each of the BMI-driven proteins on these outcomes. Two-sample MR was performed using TwoSampleMR v0.5.6 with an inverse variance weighted method and a random-effects model or Wald ratio when only one SNP was available as an instrumental variable. FDR < 0.5% was set as the threshold for significance. To minimize the risk of horizontal pleiotropy, we removed the variants associated with more than one protein in a *cis*-acting manner; therefore, we only retained the variants that were *cis*-pQTL for one protein (7008 out of 7572 variants are associated with only one protein in a *cis*-acting manner, and these 7008 variants are used as instrumental variables). To further test the absence of directional horizontal pleiotropy, we used the MR-Egger intercept test when applicable (i.e., if there are at least three instrumental variables). Additionally, we used the MR-Steiger test from TwoSampleMR v.0.5.6 to assess reverse causation, whereby cardiometabolic diseases influence plasma levels of proteins.

#### Colocalization

To ensure that the proteins and cardiometabolic diseases share the same causal genetic signal and avoid false-positive findings, We also performed colocalization using coloc R package v5.1.0^75^. We evaluated whether *cis*-pQTL of the protein shared the same causal variant with cardiometabolic diseases within 1 Mb (± 500 kb). We used default prior of *p_1_* = 10^-4^, *p2* = 10^-4^, and *p_12_* = 10^-5^ for coloc, where *p_1_* is a prior probability of trait 1 having a genetic association in the region, *p_2_* is a prior probability of trait 2 having a genetic association in the region, and *p_12_* is a prior probability of the two traits having a shared genetic association. We considered the posterior probability of a shared causal variant (PP_shared_) > 0.8 as evidence of colocalization.

#### Mediation analyses

As a validation analysis, we performed mediation analyses using network MR with a product of coefficients method. We did not adjust for the exposure (BMI) when estimating the effect of the mediator on the outcome (*β*_mediator-to-cardiometabolic_) to avoid weak instrument bias. This approach has been adopted in multiple studies^26,36-38^.

Considering that the proportion mediated can be only estimated when the direction of effects is consistent between total causal effect and causal mediation effect, we restricted the analyses to proteins that meet the following criteria: *β*_total_ × *β*_mediated_ >0

where: *β*_total_ denotes the total effect (i.e., the effect of BMI on cardiometabolic diseases), and *β*_mediated_ denotes the causal mediation effect (i.e., the effect mediated by the circulating proteins).

To estimate the causal mediation effects (*β*_mediated_), we estimated the effect of BMI on the plasma protein levels (*β*_BMI-to-protein_) and the effect of the plasma proteins on cardiometabolic diseases (*β*_protein-to-cardiometabolic_), and then multiplied these values (*β*_mediated_ = *β*_BMI-to-protein_ × *β*_protein-to-cardiometabolic_). For this, we performed MR using the same instrumental variables as in Steps 1 and 2 of MR. Subsequently, we divided *β*_mediated_ by *β*_total_ to estimate the proportion mediated and calculated the *P*-value under the null hypothesis that the protein of interest did not mediate the effect of BMI on the outcome of interest. We considered results with *P* < 0.05 to be significant. Since proteins can be correlated (e.g., in the same biological pathways), we did not apply Bonferroni correction.

### Follow-up analyses

#### Replication MR using cis-pQTL from different cohorts

To replicate the causal estimates for the effect of COL6A3 on coronary artery disease, we conducted two-sample MR using *cis*-pQTLs from different cohorts: UK Biobank^39^ (*n* = 35,571 individuals), Fenland^14^ (*n* = 10,708 individuals), and ARIC (*n* = 7,213 individuals), using the same method as described in Step 2 MR.

#### Mediation analysis with individual-level data in the EPIC-Norfolk cohort

The EPIC-Norfolk study, a component of the pan-European EPIC Study, is a cohort of 25,639 middle-aged individuals from the general population of Norfolk, a county in Eastern England^76^, who attended the baseline assessment between 1993–1998. We performed mediation analysis in a randomly selected subcohort (*n* = 872) of the EPIC-Norfolk study, in which proteomic profiling was performed using the SomaScan v4 assay. Death certificates and hospitalisation data were obtained using National Health Service numbers through linkage with the NHS digital database. Electronic health records were coded by trained nosologists according to the International Statistical Classification of Diseases and Related Health Problems, 9^th^ (ICD-9) or 10th Revision (ICD-10). Participants were identified as CAD cases if the corresponding ICD-codes (ICD-9: 410-414, ICD-10:I20-I25) were registered on the death certificate (as the underlying cause of death or as a contributing factor), or as the cause of hospitalization. The current study is based on follow-up to the 31^st^ March 2018. The case definition included all individuals identified as prevalent (at the baseline study assessment) or incident CAD cases over the follow-up period of over 20-years. The plasma protein levels were normalized with rank-based inverse normal transformation using R package RNOmni v1.01. We used the product of coefficients methods to calculate the proportion mediated, as described above, using the R package mediation^77^ v4.5.0. We used linear regression adjusting for age and sex to estimate the effect of BMI on plasma COL6A3 levels and the effect of BMI, and logistic regression adjusting for age and sex to estimate the effect of BMI on the risk of CAD and the effect of plasma COL6A3 levels on the risk of CAD. Significance of the indirect effect and the proportion mediated was estimated by computing unstandardized effects in 1000 bootstrapped samples, and calculating the corresponding 95% confidence intervals.

### Identification of the causal domain of COL6A3

#### Target region of the SomaScan v4 assay and the Olink Explore 3072 assay

We used SomaScan Menu 7K (https://menu.somalogic.com/) to determine the target amino acid sequence of two aptamers for COL6A3 from on SomaScan v4 assay with additional support from SomaLogic (Boulder, Colorado, USA). We also obtained data on the target region of Olink Explore 3072 assay from Olink (Uppsala, Sweden). In SomaScan v4 assay, two aptamers target COL6A3: one for the C-terminal of COL6A3, also known as Kunitz domain (UniProt ID: P12111, target amino acid sequence: 3108-3165) and another for the N-terminal (UniProt ID: P12111, target amino acid sequence: 26-1036). In Olink Explore 3072 assay, the assay targets the C-terminal Kunitz domain of COL6A3 with polyclonal antibody (OID20292:v1).

#### Linkage disequilibrium of COPL6A3’s cis-pQTL from the deCODE study and UK Biobank

We used the LDmatrix tool available at LDlink (https://ldlink.nci.nih.gov) with the 1000 genomes European samples as the reference panel^78^ to calculate *R*^2^ values between three SNPs: the *cis*-pQTL for COL6A3 from UK Biobank (rs1050785), the *cis*-pQTL of the C-terminal-targeting aptamer (rs11677932) from the deCODE study, and the *cis*-pQTL of the N-terminal-targeting aptamer of COL6A3 (rs2646260) from the deCODE study.

#### COL6A3 expression analyses

We downloaded bulk gene expression data in human tissues (GTEx_Analysis_2017-06-05_v8_RNASeQCv1.1.9_gene_tpm.gct.gz) from GTEx portal (https://gtexportal.org/). We generated the violin plots of *COL6A3* expression levels in each tissue using R v4.1.2. We used a two-sided Wilcoxon rank sum test to compare *COL6A3* expression in each tissue with its expression in the whole blood.

#### Single-cell RNA sequencing analysis

To investigate *COL6A3* expression at single-cell resolution in adipose tissues and coronary arteries, we reanalyzed the published expression matrix data from Emont et al.^50^ (SCP1376 at https://singlecell.broadinstitute.org/) and Wirka et al.^49^ (GSE131780 at Gene Expression Omnibus database https://www.ncbi.nlm.nih.gov/geo/), focusing on *COL6A3* expression. Following Wirka et al.^49^, we removed low-quality cells that expressed < 500 genes or had a mitochondrial content > 7.5%, and genes expressed in < 5 cells. Cells expressing > 3,500 genes were also removed to avoid bias due to doublets. The retained gene expression profiles were normalized to library size. The top 2,000 most variable genes were selected after variance-stabilizing transformation using the FindVariableFeatures function in Seurat v4.0.6. Principal component analysis was performed based on these 2,000 most variable genes after scaling and centering. Nearest-neighbor graph construction was conducted based on the first 10 principal components using the FindNeighbors function in Seurat v4.0.6 with default settings. Cell clusters were identified using the FindClusters function in Seurat v4.0.6 with default settings. Uniform Manifold Approximation and Projection (UMAP) was also performed on the first 10 principal components. Two-dimensional visualization of the cell clusters was based on the first two UMAP dimensions. We used SingleR v2.0.0 to annotate the cell clusters with the Blueprint/ENCODE dataset as the reference using default settings.

To assess whether certain cell types express *COL6A3* more significantly than others, we performed 1,000 permutations of the cell type labels and calculated the frequency (permutation p-value) of the same cell type containing the same or a larger proportion of cells expressing *COL6A3* compared to all cells.

### Follow-up analyses for the identified proteins

#### Assessment of actionability

To estimate the independent effects of fat mass and lean mass on plasma protein levels, we performed multivariable MR using fat mass and lean mass as exposures and protein levels as outcomes.

#### GWAS of fat mass and lean mass

We retrieved the GWAS data for fat mass and lean mass (i.e., fat-free mass) from UK Biobank through the OpenGWAS portal (https://gwas.mrcieu.ac.uk/). The data included 454,137 individuals of European ancestry for fat mass and 454,850 individuals for lean mass. The accession codes for the datasets were ukb-b-19393 for fat mass and ukb-b-13354 for lean mass.The fat mass and fat-free mass of the UK Biobank participants (second release, 2017) were evaluated by UK biobank with bioelectrical impedance analysis using the Tanita BC418MA body composition analyzer (Tanita, Tokyo, Japan).

#### Multivariable MR to evaluate the independent effect of fat mass and lean mass on protein levels and cardiometabolic diseases

To obtain instrumental variables, we applied the same selection criteria as in Steps 1 and 2 of MR (*P* < 5 × 10^-8^ and *r^2^* < 0.001), excluding those in the MHC region (GRCh37; chr6: 28,477,797– 33,448,354). We performed data harmonization in TwoSampleMR v0.56 and multivariable MR with the inverse variance weighted method and a random-effect model in MVMR v0.3^74^. We calculated conditional F-statistics using MVMR v0.3^74^ and evaluated whether they were above ten^73,74^ (**Supplementary Table 14 and 15**). The phenotypic correlation matrix was calculated using metaCCA v1.22.0^79^. As additional sensitivity analyses, we performed multivariable MR-Egger analysis using MendelianRandomization v0.6.085^80^.

#### Phenome-wide association study for rs11677932

We queried traits associated with the lead *cis*-pQTL of COL6A3 (rs11677932) from the deCODE study in the UK Biobank, FinGen, and GWAS catalog using the Open Target Genetics (https://genetics.opentargets.org/)

### Ethical approval

All contributing cohorts obtained ethical approval from their intuitional ethics review boards. The contributing cohorts include UK Biobank, GIANT consortium, deCODEstudy, Fenland study, AGES Reykjavik study, INTERVAL study, CARDIoGRAMplusC4D, GIGASTROKE, and MAGIC consortium. The study was approved by the Norfolk Research Ethics Committee (no. 05/ Q0101/191), and all participants gave their informed written consent.

## Data Availability

We used GWAS summary statistics from the following source:
BMI GWAS from GIANT and UK Biobank (https://portals.broadinstitute.org/collaboration/giant/).
Plasma proteome GWAS from the deCODEstudy (https://www.deCODE.com/summarydata/), UK Biobank (https://doi.org/10.1101/2022.06.17.496443), Fenland (https://omicscience.org/apps/pgwas/), and the AGES Reykjavik study (https://doi.org/.1126/science.aaq1327).
We also used coronary artery disease GWAS from CARDIoGRAMplusC4D (http://www.cardiogramplusc4d.org/), stroke GWAS from GIGASTROKE (GCST90104534 and GCST90104535, at https://www.ebi.ac.uk/gwas/studies/), and type 2 diabetes GWAS from Mahajan et al. (https://doi.org/10.1038/s41588-022-01058-3).
For gene expression data, we used data from Nathan et al. (SCP498 at Single Cell Portal https://singlecell.broadinstitute.org/) and Wirka et al (GSE131780 at Gene Expression Omnibus database https://www.ncbi.nlm.nih.gov/geo/).
Code availability
We used R v4.1.2 (https://www.r-project.org/), TwoSampleMR v.0.5.6 (https://mrcieu.github.io/TwoSampleMR/), snappy v1.0 (https://gitlab.com/richards-lab/vince.forgetta/snappy), coloc v5.1.0 (https://chr1swallace.github.io/coloc/), PLINK v1.9 (http://pngu.mgh.harvard.edu/purcell/plink/), GCTA fastGWA v1.93.3 (https://yanglab.westlake.edu.cn/software/gcta/), and Seurat v4.0.6 (https://satijalab.org/seurat/). Custom codes will be made available on GitHub (https://github.com/satoshi-yoshiji/cm_proteogenomics/) upon publication of the manuscript.

## Data availability

We used GWAS summary statistics from the following source:

BMI GWAS from GIANT and UK Biobank (https://portals.broadinstitute.org/collaboration/giant/). Plasma proteome GWAS from the deCODEstudy (https://www.deCODE.com/summarydata/), UK Biobank (https://doi.org/10.1101/2022.06.17.496443), Fenland (https://omicscience.org/apps/pgwas/), and the AGES Reykjavik study (https://doi.org/.1126/science.aaq1327).

We also used coronary artery disease GWAS from CARDIoGRAMplusC4D (http://www.cardiogramplusc4d.org/), stroke GWAS from GIGASTROKE (GCST90104534 and GCST90104535, at https://www.ebi.ac.uk/gwas/studies/), and type 2 diabetes GWAS from Mahajan *et al*. (https://doi.org/10.1038/s41588-022-01058-3).

For gene expression data, we used data from Nathan et al. (SCP498 at Single Cell Portal https://singlecell.broadinstitute.org/) and Wirka et al (GSE131780 at Gene Expression Omnibus database https://www.ncbi.nlm.nih.gov/geo/).

## Code availability

We used R v4.1.2 (https://www.r-project.org/), TwoSampleMR v.0.5.6 (https://mrcieu.github.io/TwoSampleMR/), snappy v1.0 (https://gitlab.com/richards-lab/vince.forgetta/snappy), coloc v5.1.0 (https://chr1swallace.github.io/coloc/), PLINK v1.9 (http://pngu.mgh.harvard.edu/purcell/plink/), GCTA fastGWA v1.93.3 (https://yanglab.westlake.edu.cn/software/gcta/), and Seurat v4.0.6 (https://satijalab.org/seurat/). Custom codes will be made available on GitHub (https://github.com/satoshi-yoshiji/cm_proteogenomics/) upon publication of the manuscript.

## Acknowledgments

The Richards research group is supported by the Canadian Institutes of Health Research (CIHR: 365825, 409511, 100558, 169303), the McGill Interdisciplinary Initiative in Infection and Immunity (MI4), the Lady Davis Institute of the Jewish General Hospital, the Jewish General Hospital Foundation, the Canadian Foundation for Innovation, the NIH Foundation, Cancer Research UK, Genome Québec, the Public Health Agency of Canada, McGill University, Cancer Research UK [grant number C18281/A29019] and the Fonds de Recherche Québec Santé (FRQS). J.B.R. is supported by an FRQS Mérite Clinical Research Scholarship. Support from Calcul Québec and Compute Canada is acknowledged. TwinsUK is funded by the Welcome Trust, Medical Research Council, European Union, the National Institute for Health Research (NIHR)-funded BioResource, Clinical Research Facility and Biomedical Research Centre based at Guy’s and St Thomas’ NHS Foundation Trust in partnership with King’s College London. NJT is a Wellcome Trust Investigator (202802/Z/16/Z), is the PI of the Avon Longitudinal Study of Parents and Children (MRC & WT 217065/Z/19/Z), is supported by the University of Bristol NIHR

Biomedical Research Centre (BRC-1215-2001), the MRC Integrative Epidemiology Unit (MC_UU_00011/1) and works within the CRUK Integrative Cancer Epidemiology Programme (C18281/A29019).

The Genotype-Tissue Expression (GTEx) Project was supported by the Common Fund of the Office of the Director of the National Institutes of Health, and by NCI, NHGRI, NHLBI, NIDA, NIMH, and NINDS. The data used for the analyses described in this manuscript were obtained from: the GTEx Portal on March 26, 2023.

S.Y. is supported by the Japan Society for the Promotion of Science. T.L. is supported by a Schmidt AI in Science Postdoctoral Fellowship, a Vanier Canada Graduate Scholarship, an FRQS doctoral training fellowship, and a McGill University Faculty of Medicine Studentship. G.B.L. is supported by scholarships from the FRQS, the CIHR, and Québec’s ministry of health and social services. Y.C. is supported by an FRQS doctoral training fellowship and the Lady Davis Institute/TD Bank Studentship Award. The funders had no role in study design, data collection and analysis, decision to publish, or preparation of the manuscript. We acknowledge Biorender (biorender.com) for providing materials used to create the illustrative diagram.

---

**Extended Figure 1.**
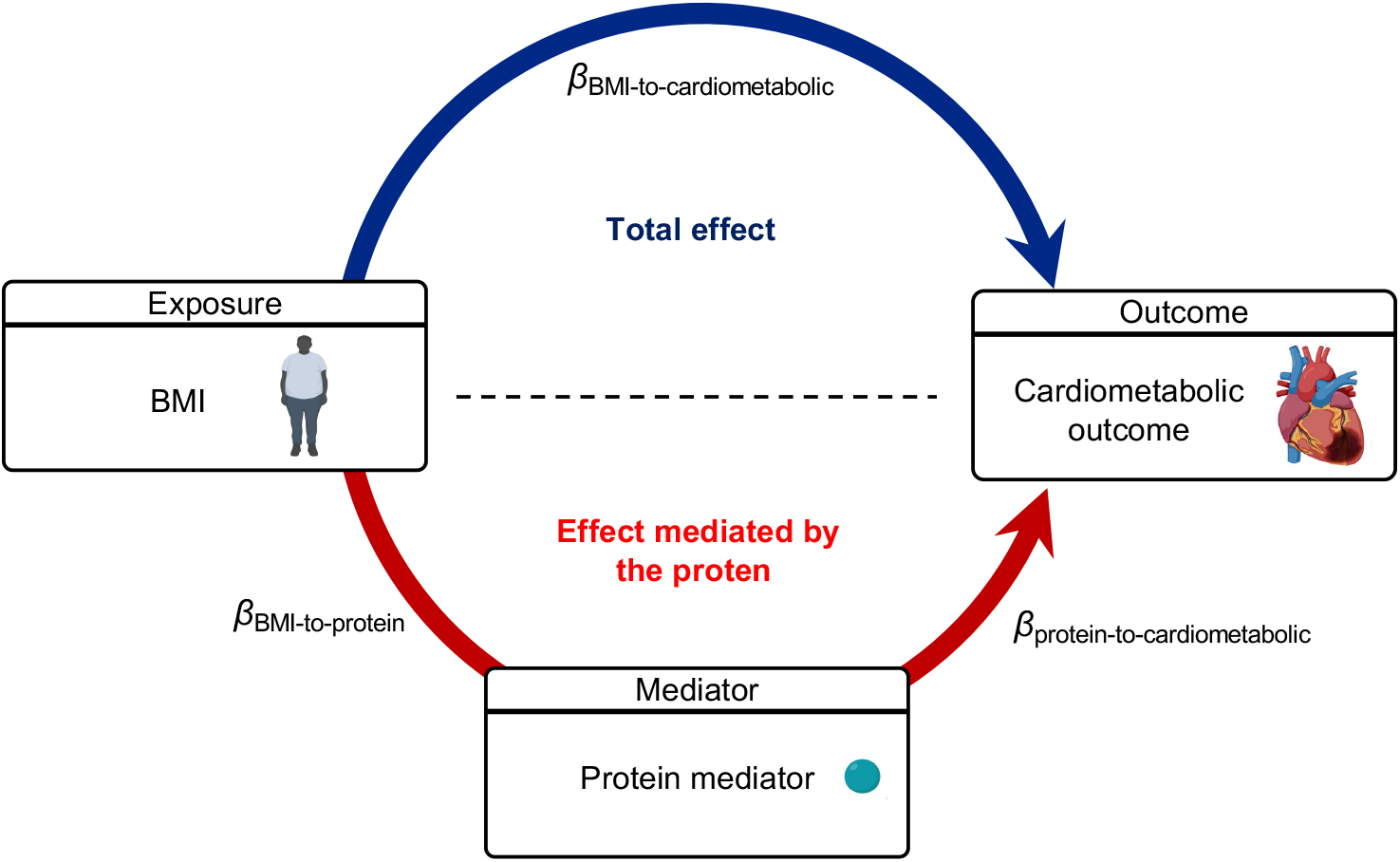
Schematic illustration of the mediation analysis. The figure demonstrates the causal relationship between BMI, the protein mediator, and cardiometabolic diseases using directed acyclic graphs. The dark blue arrow represents the total effect of BMI on cardiometabolic diseases (*β*_BMI-to-cardiometabolic_), while the red arrow represents the effect of BMI on cardiometabolic diseases mediated by the protein mediator. To calculate the ratio mediated, we used the product of coefficients method. This involved multiplying the effect of BMI on the protein mediator (*β*_BMI-to-protein_) by the effect of the protein mediator on cardiometabolic diseases (*β*_protein-to-cardiometabolic_) to estimate the effect mediated by the protein (*β*_mediated_ = *β*_BMI-to-protein_ × *β*_protein-to-cardiometabolic_). Subsequently, we divided *β*_mediated_ by *β*_total_ to estimate the proportion mediated and calculated the *P*-value under the null hypothesis that the protein of interest did not mediate the effect of BMI on the outcome of interest. BMI: body mass index, MR: Mendelian randomization.

